# A Randomised, Multi-centre Phase II Trial of Weekly Paclitaxel and Vistusertib in Platinum-Resistant Ovarian High-Grade Serous Carcinoma: OCTOPUS Arm 1

**DOI:** 10.1101/2022.05.26.22275618

**Authors:** S. Banerjee, G. Giannone, A. Clamp, D. Ennis, R. Glasspool, R. Herbertson, J. Krell, R. Riisnaes, H.B. Mirza, Z. Cheng, J. McDermott, C. Green, R.S. Kristeleit, A. George, C. Gourley, L-A Lewsley, D. Rai, U. Banerji, S. Hinsley, I.A. McNeish

## Abstract

**Background:** Preclinical studies support targeting PI3K/AKT/mTOR signalling in platinum-resistant ovarian cancer (PROC). A phase I study of the dual mTORC1/mTORC2 inhibitor vistusertib with weekly paclitaxel (wP) showed activity. We report the results of Arm 1 of OCTOPUS, the first randomised trial of weekly paclitaxel and dual mTORC1/2 inhibition in ovarian cancer.

**Methods:** Patients with platinum-resistant or refractory high grade serous carcinoma were randomised (1:1) to wP (80mg/m^2^ D1,8,15 of 28 day cycle) plus oral vistusertib (50mg BD) or placebo (P). The primary endpoint was progression-free survival (PFS). Secondary endpoints included response rate (RR) and overall survival (OS).

**Results:** 140 patients (median age 63, range: 36-86; 18% platinum-refractory; 54% ≥3 prior therapies) were randomised. There was no difference in PFS (median 4.5 vs 4.1m (HR 0.84; 80% CI (0.67, 1.07); 1-sided p=0.18), OS (median 9.7 vs 11.1m (HR 1.21; 80% CI (0.91, 1.60); 1-sided p=0.80) or RR (odds ratio 0.86; 80% CI (0.55, 1.36); 1-sided p=0.66). Grade 3/4 adverse events were 41.2% (wP+V) vs 36.7% (wP+P). Low tumour PTEN expression was associated with longer PFS in the wP+V arm (9.4 vs 4.1m p=0.003) but not in the wP arm (4.8 vs 4.2m p=0.60). Tumour genome-wide copy number (CN) analysis suggested that high CN signature 4 was associated with worse outcome in the wP+P arm (2.3 vs 4.6m p=0.018) but not the wP+V arm (5.4 vs 3.3m).

**Conclusions:** Vistusertib did not improve clinical activity of wP in PROC. However, low tumour cell PTEN expression may be a predictive biomarker for vistusertib activity.

**Translational Relevance:** Preclinical studies suggest that activation of the PI3K/AKT/mTOR signalling pathway contributes to platinum-resistance in ovarian high grade serous carcinoma (HGSC). Based on activity in a phase I study, we evaluated the clinical efficacy of the dual mTORC1/mTORC2 inhibitor vistusertib in combination with weekly paclitaxel in the OCTOPUS study - a multi-centre, randomised, placebo-controlled, phase II trial in platinum-resistant ovarian (HGSC). In the first randomised trial of weekly paclitaxel and dual mTORC1/2 inhibition in ovarian cancer, vistusertib did not improve clinical activity of weekly paclitaxel. However, translational analyses indicated that low tumour cell PTEN expression may be a predictive biomarker for vistusertib activity. We also showed genome-wide copy number (CN) analysis, in particular high exposure to CN signature 4, may also allow identification of patients with greater chance of benefit from dual mTORC inhibition. Potential predictive biomarkers identified in our study should be evaluated in ongoing/future studies.

## Introduction

Despite improvements in first line treatment for advanced ovarian cancer, approximately 85% of patients recur and eventually develop fatal chemotherapy resistance. The duration of response following platinum therapy (platinum-free interval) remains an important consideration when selecting treatment options for recurrent ovarian cancer. For patients whose disease has relapsed during (platinum-refractory) or within 6 months of completing platinum-based therapy (‘platinum-resistant’), treatment options are limited. Weekly paclitaxel has activity in platinum-resistant ovarian cancer, with response rates of approximately 30%. However, the median PFS is generally 3-4 months and overall survival approximately 12 months (1).

One potential mechanism of resistance to both platinum and taxane chemotherapy is activation of the PI3K/AKT/mTOR signalling pathway (2,3). Raised p-S6K expression in malignant cells isolated from ascites in ovarian cancer patients has been associated with chemoresistance and poor clinical outcome (4). However, response rates in single arm studies of first generation mTOR inhibitors such as temsirolimus were poor (5), which prompted the development of dual mTORC1/2 inhibitors that could prevent the AKT-mediated feedback observed with these rapalogues. Following encouraging preclinical data (6), a phase I study of weekly paclitaxel in combination with the dual mTORC1/2 inhibitor vistusertib was conducted with a dose expansion for patients with ovarian high grade serous carcinoma (HGSC). The recommended phase II dose (RP2D) was established as 80 mg/m^2^ paclitaxel with 50mg vistusertib bd 3 days per week. The RECIST response rate in the HGSC cohort was 52% (13/25), with median PFS of 5.8 months (95% CI: 3.3–18.5) (7). Here, we evaluated the clinical efficacy of vistusertib in combination with weekly paclitaxel in the OCTOPUS study - a multi-centre, randomised, placebo-controlled, phase II trial in platinum-resistant ovarian high grade serous carcinoma.

## Methods

### Study design

OCTOPUS was a phase 2, randomised, double-blind, placebo-controlled, multicentre study evaluating the efficacy and safety of vistusertib in combination with paclitaxel compared to paclitaxel alone in patients with high grade serous ovarian carcinoma (HGSC) progressed during (platinum-refractory) or within 6 months of completing last platinum-based therapy (platinum-resistant). The study was conducted in accordance with the Research Governance Framework for Health and Community Care (Second edition; 2006) and the Medicines for Human Use (Clinical Trials) Regulations, 2004 SI 2004:1031 (as amended) and World Medical Association Declaration of Helsinki Ethical Principles for Medical Research Involving Human Subjects 1964 (as amended) and was co-sponsored by University of Glasgow and NHS Greater Glasgow and Clyde. Ethical approval was obtained from London–Brighton and Sussex Research Ethics Committee (reference 15/LO/1302) and all patients provided written informed consent.

### Patient population

Patients (≥18 years) with histologically confirmed high grade serous carcinoma of ovarian, fallopian tube or primary peritoneal origin, with relapse in the platinum-resistant timeframe, were enrolled. Platinum resistant/refractory status was defined as either radiological progression (based on RECISTv.1.1) or CA125 rise (according to GCIG CA125 criteria) plus symptoms indicative of progression either during (platinum-refractory) or within 6 months of completing (platinum-resistant) prior platinum therapy. Measurable or evaluable disease according to RECISTv.1.1 and/or GCIG CA125 criteria was required (8). Patients who received weekly paclitaxel in combination with platinum as part of first line treatment were eligible if the interval since the last dose of weekly paclitaxel was >6 months at the time of randomisation. Patients who received prior weekly paclitaxel for platinum-resistant disease were excluded. A biopsy was mandatory at study entry if deemed technically feasible. There were no restrictions on number of lines of prior therapy (see supplementary for full inclusion/exclusion criteria) and the most recent chemotherapy did not have to be platinum-based.

### Study Design, Treatment and Conduct

140 participants were randomised (1:1) to receive treatment as follows:

- wP+V: paclitaxel 80mg/m^2^ IV D1, 8, 15 plus Vistusertib 50mg bd days 1-3, 8-10 and 15-17 of a 28 day cycle (experimental arm)
- wP+P: paclitaxel 80mg/m^2^ IV D1, 8, 15 plus placebo bd days 1-3, 8-10 and 15-17 of a 28 day cycle (control arm)

Patients received 6 cycles (24 weeks) of combination treatment. Thereafter, patients who did not have progressive disease and had completed at least 4 cycles of combination treatment could continue on vistusertib (50mg bd)/placebo alone as continuous maintenance therapy. Patients could continue beyond 6 cycles of paclitaxel and vistusertib/placebo before receiving maintenance therapy, at the discretion of the Investigator and Chief Investigator, provided the patient had not progressed.

After informed consent and eligibility confirmation, participants were allocated to treatment using minimisation with a random element, and the following minimisation factors: treatment centre, measurable disease (yes vs. no), platinum refractory vs. resistant, and taxane-free interval (<6 months vs. ≥6 months vs. no prior taxane) (1).

### Assessments

Imaging assessments (CT chest, abdomen and pelvis or MRI) were performed at baseline and then 8 weekly until progression. CA125 was measured 4 weekly. Adverse events were graded according to National Cancer Institute Common Terminology Criteria for Adverse Events (NCI-CTCAE) v4.3. Quality of life (QoL) was assessed using EQ-5D at baseline, prior to each cycle of treatment, end of treatment and follow up.

### Statistical Considerations

The primary end point was progression-free survival (PFS) in the intention-to-treat (ITT) population, defined as the time from randomisation until the first appearance of confirmed progressive disease as defined by a combined RECIST v1.1 and GCIG CA125 criteria (8) or death from any cause. Patients still alive and without progression at the time of analysis were censored at the date last known to be alive and progression-free. Secondary endpoints were Response rate (RR) (best recorded response according to combined RECISTv1.1 and GCIG CA125 criteria), Overall Survival (OS), defined as the time from randomisation until death from any cause, safety and tolerability (according to NCI CTCAE Version 4) and Quality of Life as measured by EQ-5D.

The trial followed a three-outcome design (9): a PFS difference in favour of wP+V that was statistically significant at 10% was a clear signal that a subsequent phase III study is warranted. A result statistically significant at the 20% level (but not 10%) would require supportive data with improved response rate before a subsequent phase III would be considered.

A total of 122 PFS events were required to detect a 50% improvement in median PFS from 3.7 months with wP+P to 5.55 months with wP+V with 90% power, at the 20% 1-sided level of statistical significance (or equivalently with 80% power at the 10% level of statistical significance). This required 140 patients (70 patients per arm) recruited over 16.5 months with 8 months subsequent follow-up. This incorporated an interim analysis for futility, after 40 PFS events had been observed, using a Lan-DeMets spending function (10) and Pocock type boundary (11).

All analyses were pre-planned, unless specified, and performed using SAS Enterprise Guide 5.1. Efficacy analyses were undertaken on the ITT population, including all patients randomised into the study. Safety and tolerability analyses were undertaken on the safety population, defined as participant who received at least one dose of study treatment.

PFS and OS were analysed using Cox regression via a model incorporating the blinded study arm and the factors used in the minimisation algorithm. A test of the proportional hazards (PH) assumption was performed by testing the significance of a time dependent covariate in the Cox model, as well as being assessed graphically. Response rates (complete and partial combined) were compared between the blinded study arms using logistic regression via a model incorporating study arm and the factors used in the minimization algorithm.

QoL was analysed using mixed effects and repeated measures models, adjusting for minimisation factors, as well as time-point and interaction terms (a change from the SAP). Both complete case analyses (CC; ignoring missing data), and analyses accounting for missing values using multiple imputation (MI) with 40 imputations were undertaken (12).

#### Translational analyses

Details of the translational samples and analysis are given in Supplementary Methods.

## Results

### Patient Demographics

A total of 140 patients were randomised (n=70 vistusertib, n=70 placebo) from 20 UK sites between January 2016 and March 2018 and included in the ITT population (Figure 1). Baseline characteristics are summarised in Table 1. The median age was 65 years (IQR 58–70) in the wP+V arm and 61 years (58-66) in the wP+P arm. 51% and 55% respectively had ≥3 prior treatment lines; 9% and 6% respectively had a taxane-free interval of <6 months whilst 29 patients (20.7%) had received prior weekly paclitaxel in combination with carboplatin, two of whom had received it twice. 139/140 patients had high grade serous histology (1 patient deemed ineligible after starting trial treatment was confirmed to have low grade serous carcinoma) and 17.9% were platinum-refractory.

**Table 1.** Baseline characteristics

|  |  | **wP+V**<br>**(n=70)** | **wP+P**<br>**(n=70)** | **Total**<br>**(n=140)** |
| --- | --- | --- | --- | --- |
| **RECIST**<br>**Measurable**<br>**disease** | Yes | 62 (88.6%) | 61 (87.1%) | 123 (87.9%) |
|  | No | 8 (11.4%) | 9 (12.9%) | 17 (12.1%) |
| **Platinum**<br>**status** | Refractory (progressed on platinum) | 12 (17.1%) | 13 (18.6%) | 25 (17.9%) |
|  | Resistant (progressed within 6 months of completing platinum) | 58 (82.9%) | 57 (81.4%) | 115 (82.1%) |
| **Taxane-free**<br>**interval** | <6 months | 6 (8.6%) | 4 (5.7%) | 10 (7.1%) |
|  | ≥6 months | 58 (82.9%) | 64 (91.4%) | 122 (87.1%) |
|  | No Prior Taxane | 6 (8.6%) | 2 (2.9%) | 8 (5.7%) |
| **Age** | Median (IQR) | 65.0 (58–70) | 61.0 (58–67) | 63.0 (58–69) |

**Figure 1.**
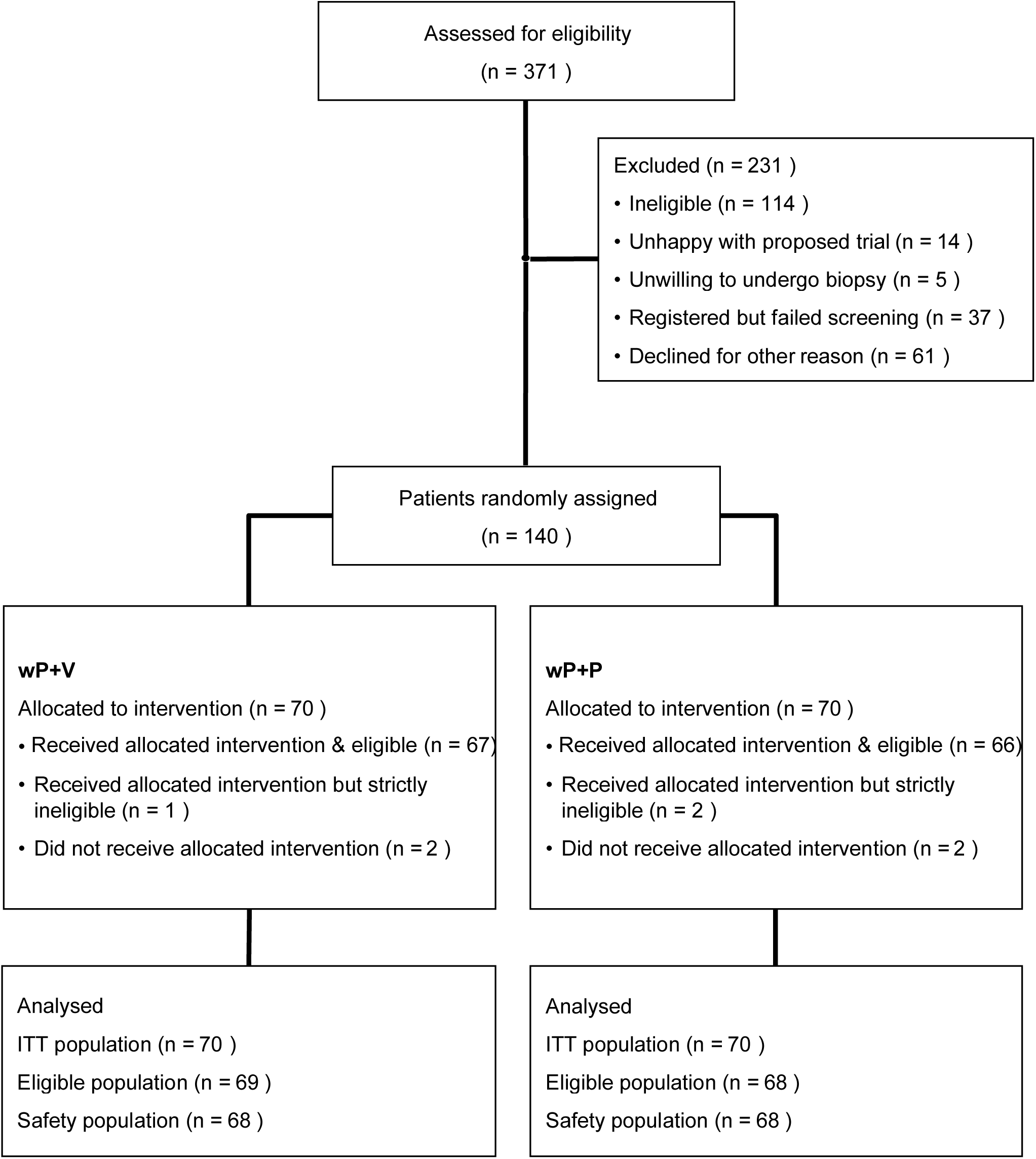
CONSORT diagram

### Efficacy

Median PFS was 4.5 (wP+V) vs 4.1 months (wP+P) (adjusted hazard ratio (HR) 0.84; 80% CI (0.67, 1.07); 1-sided p=0.18) (Figure 2A). Median OS was 9.7 vs 11.1 months (adjusted HR 1.21; 80% CI (0.91, 1.60); 2-sided p=0.80) (Figure 2B). Testing of the proportional hazards assumption confirmed that the above hazard ratios from the Cox proportional hazards model are appropriate.

**Figure 2A.**
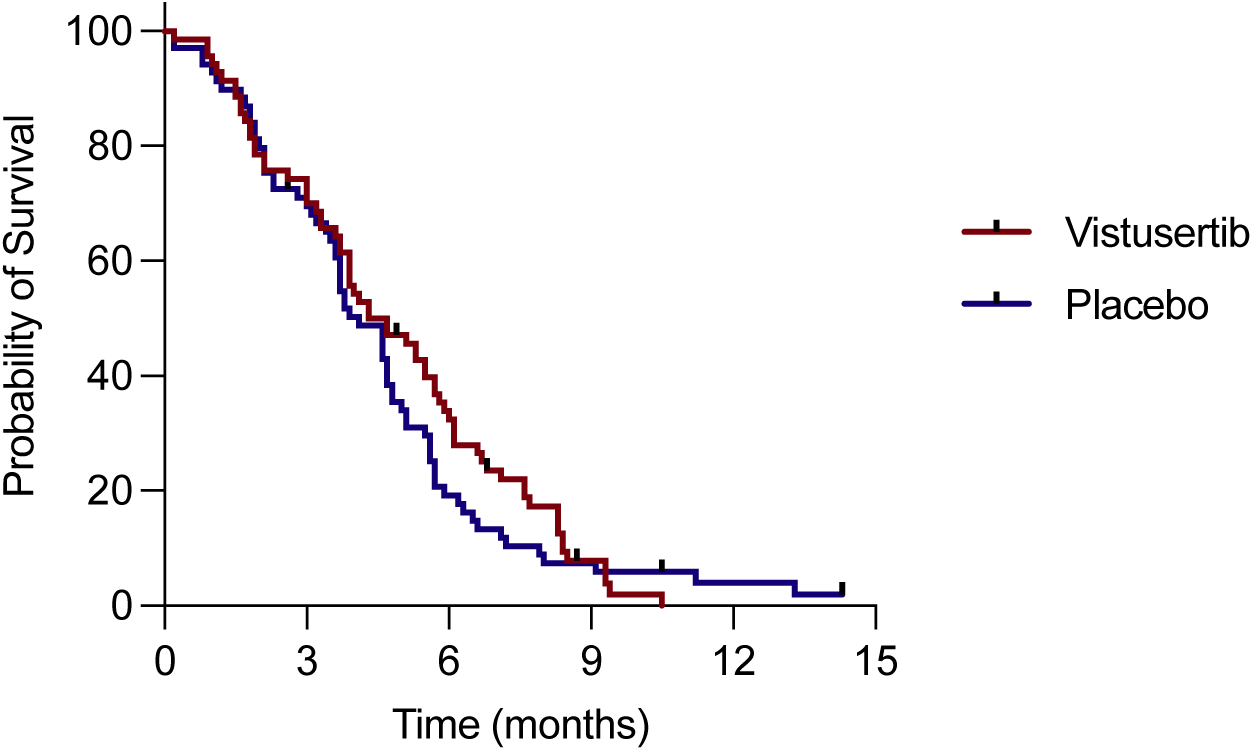
Progression-free survival: Kaplan-Meier curve

**Figure 2B.**
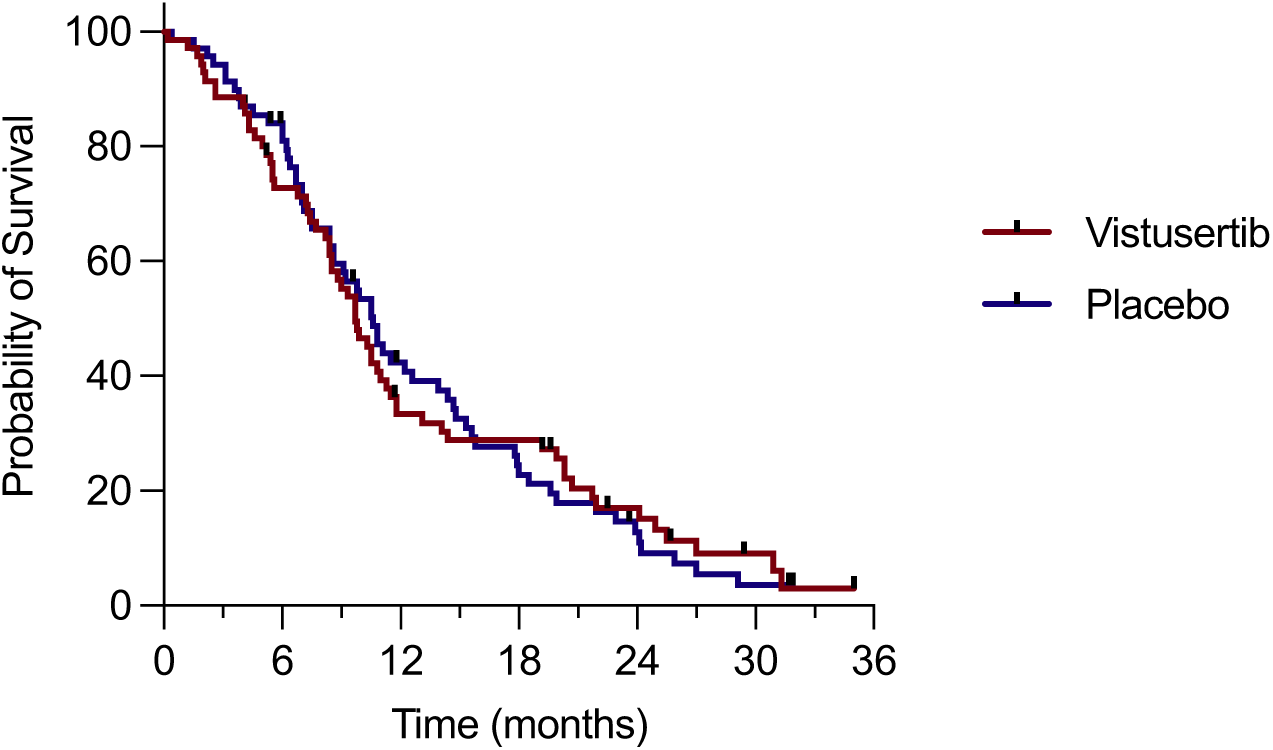
Overall survival: Kaplan-Meier curve

The RECIST 1.1 response rates were 29% vs 30% for wP+V and wP+P respectively and GCIG combined RECIST 1.1/CA125 criteria response rates were 53% and 54% respectively. There was no difference in response rate by combined GCIG RECIST1.1/CA125 criteria (adjusted odds ratio 0.86; 80% CI (0.55, 1.36); 1-sided p=0.66). Although PFS was significant at the 20% level, there was no improvement in response rate and therefore the primary endpoint was not met as per statistical considerations.

The Forest plot in Figure 2C indicates the estimated hazard ratios for clinical minimisation factors with p-values for interaction test. There was no difference in PFS or OS according to measurable disease status or platinum status (refractory/resistant). Although there appears to be an interaction with taxane-free interval, a further analysis splitting taxane-free interval into approximate quartiles (to account for bias due the small number of patients in the <6 months group) showed no evidence of an interaction (p=0.83).

**Figure 2C.**
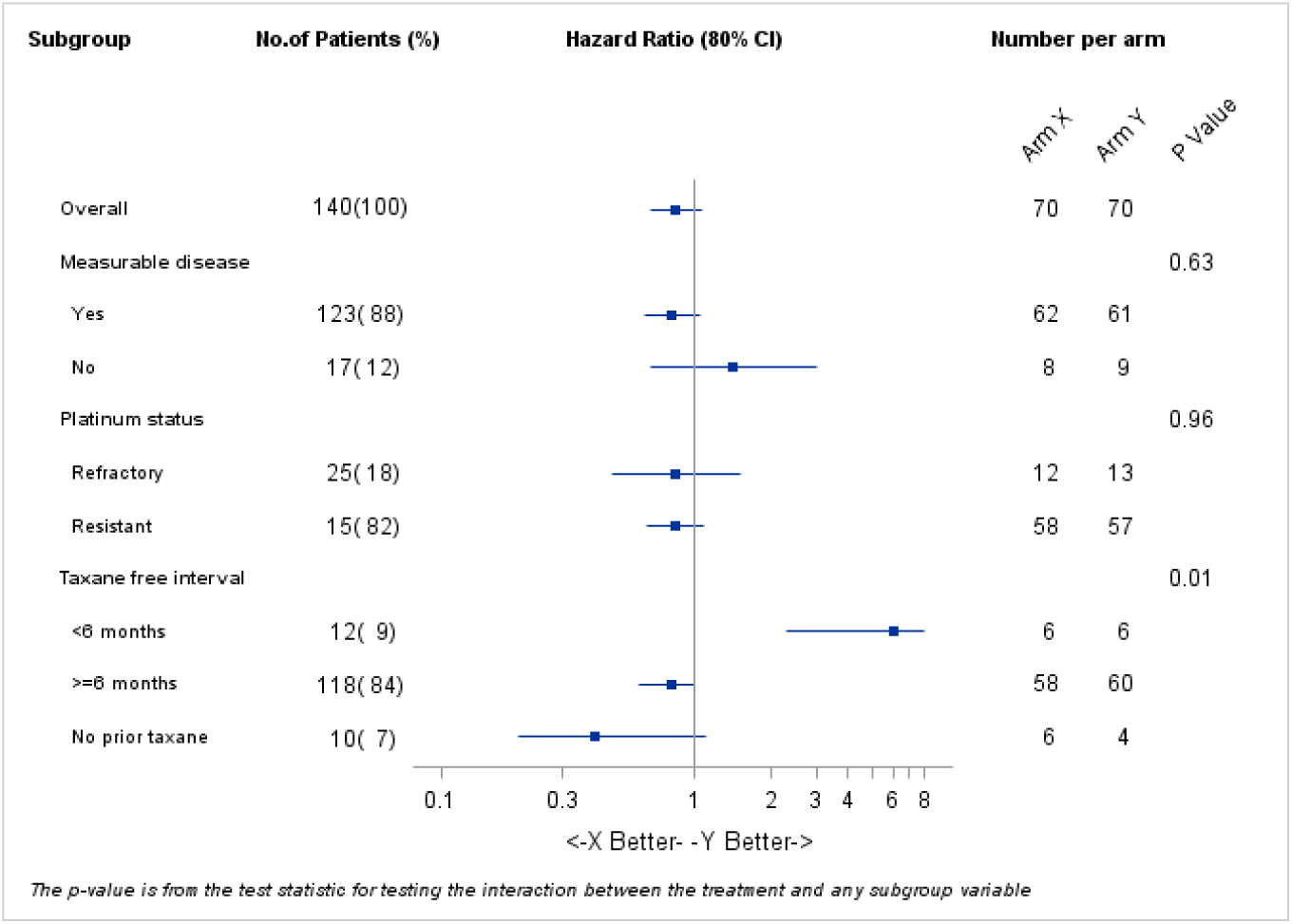
Forest plot for clinical minimisation factors with p-values for interaction test

### Tolerability

136 participants were included in the safety population (n=68 in each arm). Table S1 summarises adverse events that occurred during combination therapy at grade 2 or above in >10% of patients in either arm or that are different significantly between the arms. Grade 3/4 adverse events were reported in 41.2% (wP+V) vs 36.7% (wP+P). Grade 1/2 adverse events were reported in 58.9% vs 63.2%. The most frequent grade 3/4 toxicities were lymphopenia (13% wP+V vs 9% wP+P) and fatigue (9% vs 4%). There was significantly more gastro-oesophageal reflux (grade 1/2 10% v 0%), rash (grade 2/3 9% v 0%) and lymphopenia (grade 2/3/4 47 v 31%) with wP+V compared to wP+P. There were no grade 4 or 5 events.

### Treatment delivery

Table S2 summarises the treatment delivery of weekly paclitaxel and vistusertib/placebo. Dose intensity of weekly paclitaxel was similar between the wP+V and wP+P arms. Dose reductions and missed doses were greater for vistusertib than placebo (>1 dose reduction 19% vs 6%). The rate of discontinuation of vistusertib or placebo due to toxicity was 5% vs 0%.

### Quality of Life

126 patients were included in the QoL analysis. HRQoL questionnaires were completed at baseline (94.3%), weekly during treatment and 8-weekly post treatment. Overall QoL completion at time-points used in the analysis (up to and including week 28) was 79.3%. Figure S1 shows the EQ5D scores by time point. There were no statistically significant differences (at the 10% or 20% level) between the two treatment arms for EQ5D index or EQ5D VAS (p>0.25 in all analyses).

### Biomarker Analysis

Digital quantification of PTEN status showed a high correlation between QuPath and pathologist scores (r=0.94, *p*<0.0001 for tumour: r=0.70, *p*=0.009 for non-tumour – Figure S2, S3). Expression of PTEN was more variable in tumour cells (H-score range 23-258) than in non-tumour cells (range 44-177). Using pre-defined criteria (see Supplementary Methods), 13 (19.1%) cases were defined as PTEN low and 55 (80.9%) as PTEN high. There was a significant interaction between the randomisation arm and PTEN status (*p*=0.015) for PFS. Patients with PTEN low tumours showed a longer PFS compared with PTEN high patients in the experimental arm (median 9.4 (95%CI 2.8-16) vs 4.1 (95%CI 1.1-7) months respectively *p*=0.009; HR =0.14 (95%CI 0.03-0.62) but not in the control arm (4.8 (95%CI 2.5-7.0) vs 4.2 (95%CI 2.5-5.9) months respectively *p*=0.60) (Figure 3). No difference in OS was recorded although there was a trend toward a longer OS for PTEN low patients in the experimental arm (p=0.076).

**Figure 3.**
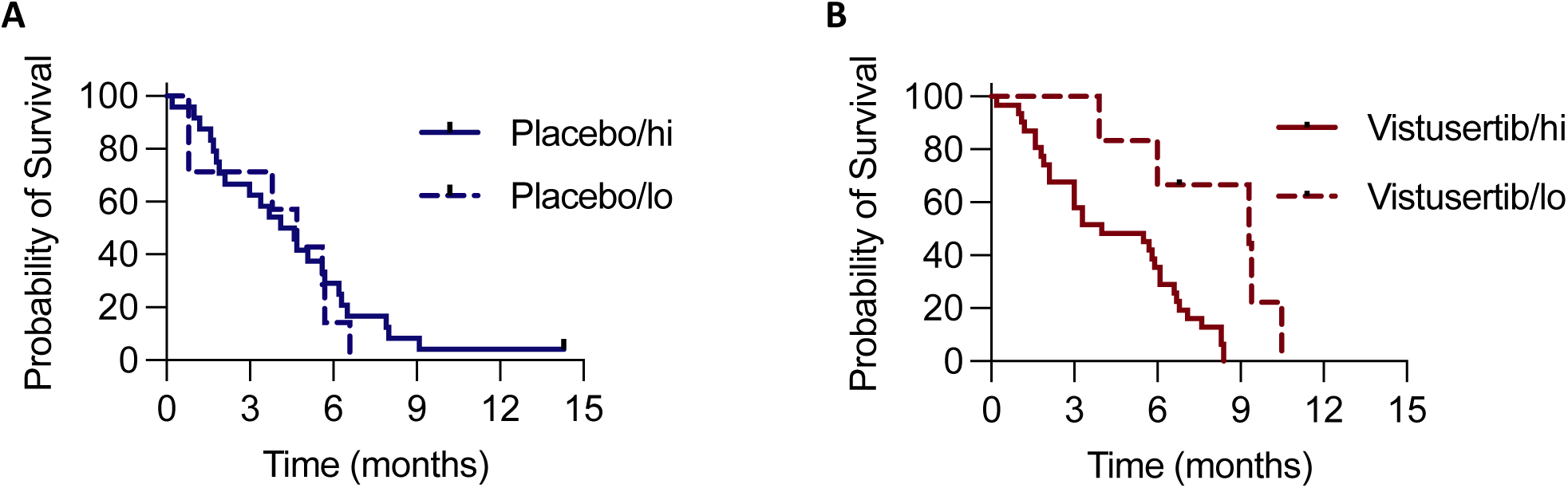
Progression-free survival according to PTEN IHC status A: Placebo; median PFS 4.4m (PTEN high) vs 4.7m (PTEN low); p=0.6077 B: Vistusertib: median PFS 4.0m (PTEN high) vs 9.3m (PTEN low); p=0.0031 HR=0.31 (95%CI 0.16–0.63)

We used sWGS to analyse genome-wide absolute CN in 78 samples from 66 patients (with 12 matched study entry and archival pairs). There were no differences in ploidy, rates of focal somatic CN alterations or CN signature exposure between diagnosis and study entry (Figure S4), allowing us to combine archival and study entry samples as a single cohort for survival analyses. No CN signature had a statistically significant predictive value although a trend toward an interaction between the treatment and exposure to normalized signature 4 (*p*=0.071) was recorded for PFS. Therefore, we divided our cohort in two groups according to a pre-defined cut-off, the mean normalized signature 4 exposure. 40 (60.6%) patients had high exposure (defined as ≥mean) and 26 (39.4%) low (defined as <mean). A high exposure to normalized signature 4 was associated with a significantly worse outcome in the control arm with a PFS of 2.3 (95%CI 0.2-4.4) vs 4.6 (95%CI 3.1vs 6.2) months respectively, *p*=0.018 but a numerically longer PFS of 5.4 (95%CI 2.7-8.1) vs 3.3 (95%CI 1.7-4-9) months respectively, *p*=0.125 in the experimental arm (Figure 4). However, there was no statistical difference in OS in the experimental or the control arm according to the exposure to normalized signature 4.

**Figure 4:**
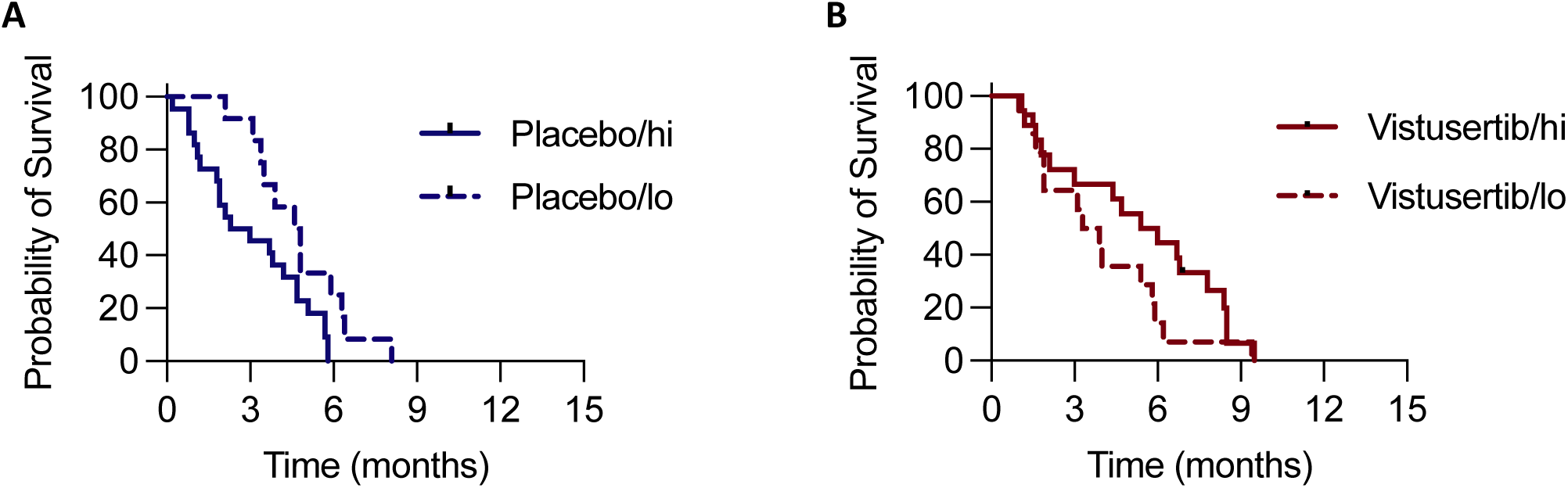
PFS according to copy number signature 4 (CN4) exposure A: Placebo; median PFS 2.7m (CN4 high) vs 4.7m (CN4 low); p=0.02 HR=0.47 (95%CI 0.24–0.93) B: Vistusertib: median PFS 5.7m (CN4 high) vs 3.6m (PTEN low); p=0.11

## Discussion

Recurrent ovarian cancer that is resistant or refractory to platinum-based chemotherapy has a dismal prognosis and there have been strikingly few positive randomised clinical trials in this patient population (13,14). Treatment still relies upon single agent chemotherapy, usually weekly paclitaxel or pegylated liposomal doxorubicin and less commonly topotecan, with response rates that rarely exceed 30% (0-30%) (15). Previous attempts to improve activity of weekly paclitaxel by targeting the insulin, SRC, HER2 and MAPK signalling pathways have been largely negative (1,16-19). The key exception is targeting angiogenesis using bevacizumab (15), although the activity of the weekly paclitaxel/bevacizumab combination was reported in an exploratory, sub-group analysis. Moreover, many patients receive bevacizumab in the first-line or platinum-sensitive relapsed setting (20-22), precluding its use in the platinum-resistant setting. In addition, the use of bevacizumab in platinum-resistant disease is limited to two or fewer lines of therapy and excludes women with platinum-refractory disease or bulky disease on the bowel serosa (23,24).

Abnormalities in the PI3K/AKT pathway are frequent in HGSC, most commonly due to amplifications or activating mutations in *PIK3CA*, loss of *PTEN* through deletion or genomic rearrangements and amplification of *AKT1/2* (25,26), and may be key drivers of drug resistance (27). However, translating this knowledge into effective therapies has proven extremely challenging (28) and predictive biomarkers of response in HGSC and other cancers remain elusive (29).

We believe that OCTOPUS is the first randomised trial to evaluate whether the addition of a dual mTORC inhibitor to weekly paclitaxel might improve PFS and OS in platinum resistant HGSC. We also believe that this is the first platinum-resistant/refractory study exclusively in the HGSC histological subtype and the first to permit prior weekly paclitaxel treatment (if more than 6 months from study entry). Of note, 18% of patients had platinum-refractory disease and 20.7% had received prior weekly paclitaxel in combination with carboplatin. The combination of vistusertib and weekly paclitaxel showed promise in an expanded HGSC phase I cohort, with radiological responses seen in 13/25 (52%) patients and the median PFS was 5.8 months (7). Although in this randomised phase II trial, the safety profile was manageable and the addition of vistusertib did not impair QoL compared to the control arm, the study failed to reach its primary endpoint. The observed outcomes in the control arm (ORR 30%, median PFS 4.4 months; median OS 11.1 months) were in keeping with previous studies of weekly paclitaxel in platinum-resistant ovarian cancer (1,16) and there was a small improvement in PFS (HR=0.84) in the experimental arm. However, this was not accompanied by improved response rate. Moreover, OS, the key endpoint in randomised trials in this population (30), showed no improvement in the experimental arm.

This study is the first arm of OCTOPUS, a novel phase II umbrella framework that aims to test the addition of new target drugs to weekly paclitaxel, comparing these combinations to a rolling control arm with paclitaxel alone. OCTOPUS provides a flexible and rapid platform to move promising combinations from phase I into a randomised assessment. The current study emphasises the importance of randomised trials and also the importance of collecting archival and study entry samples and embedding biomarker research embedded within clinical trials.

The search for robust and reliable biomarkers of response to PI3K/AKT/mTORC inhibition has repeatedly shown that assessment of single nucleotide variants in individual genes in the pathway is not predictive (7). A recent meta-analysis of 233 ovarian cancer patients enrolled into 19 studies of single agent PI3K/AKT/mTORC pathway inhibitors showed an overall response rate of 3% (31) with no improvement in observed clinical benefit rate in studies that selected patients based upon current biomarkers, including assessment of single gene status using next generation sequencing. Certainly, a recent analysis of PI3K/AKT/mTORC signalling in ER+ breast cancer, as assessed by gene expression, found no correlation with *PI3KCA* mutation status (32). Moreover, there is an increasing understanding of the importance of PI3K/AKT/mTORC pathway signalling in the tumour microenvironment in cancer, including the differentiation and activity of both innate and adaptive immune cell populations (33). Thus, newer biomarkers will need to assess both tumour and non-tumour cells and provide readouts of overall pathway activity.

Here we utilised two potential predictive biomarkers. Digital image analysis allows automated segmentation of tumour vs non-tumour cells and produces reproducible and quantitative assessment of protein expression. Given the key role of PTEN in regulating PI3K/AKT/mTORC signalling and the frequency PTEN loss in HGSC (34), we assessed PTEN expression by IHC using validated protocols. Previous results confirm that stromal PTEN expression significantly contributes to overall expression within HGSC (35), reinforcing the importance of separate assessment of tumour and non-tumour cells. Using pre-defined classification, our results indicate that low tumour cell PTEN expression is associated with improved PFS in the experimental arm but, importantly, not in the control arm. There are conflicting data on the prognostic importance of PTEN loss – initial studies (34) suggested a negative effect on survival, but this was not confirmed in a largescale analysis by the OTTA consortium (35). Certainly, our analysis of archival material using the same protocols as OTTA appear to confirm that PTEN loss is not simply a prognostic biomarker. We believe that this is one of the first uses of digital pathological quantification in ovarian cancer trials, but any predictive effect of PTEN loss will obviously need to be confirmed prospectively in a separate study cohort. The on-going DICE trial (weekly paclitaxel +/− TAK-228 clinicaltrials.gov NCT03648489) will provide such an opportunity.

HGSC is the archetypal C class tumour (36), driven very largely by copy number abnormalities rather than mutations (SNV or indels). We recently described copy number signatures, patterns of copy number alterations in HGSC that reflected underlying mutational processes (37). Thus, analysis of CN signatures may reveal the activity of key driver processes in specific cancers. CN signature 4, marked by high segment copy number, was significantly associated with whole genome duplication and mutations in the PI3K/AKT pathway (37). Our recent comparison of early- and late-stage HGSC genomes confirmed the link between CN signature 4 and high ploidy, as well as its association with poor prognosis (38). Here, we showed that high CN signature 4 exposure was again negatively prognostic in the control arm but was associated with an improvement in PFS in the experimental arm, although this did not reach statistical significance.

There are several shortcomings in this study. The first and foremost is that it failed to reach its pre-defined primary endpoint. Our translational biomarkers, although pre-defined at the start of the translational studies, were not pre-planned and were used neither as stratification factors nor to select patients for study entry. In addition, the number of samples available for the translational analyses was low, despite provision of archival FFPE material being mandatory. Moreover, we used simple cut-offs to define PTEN high vs low and CN signature 4 high vs low, and although such delineators have been used previously, they are pragmatic rather than biological. Both protein expression and CN signature exposure are continuous variables and application of cut-offs should ideally be based on biologically defined thresholds. The inability of current HRD assays to identify populations of patients who do not benefit from PARP inhibitor maintenance treatment is a stark reminder of the challenges of categorising continuous variable biological data (39,40). A further challenge with CN signature analysis is that the data are compositional (ie sum to 1 in each sample) meaning that any increase in one signature must, by definition, by accompanied by a decrease in at least one other; thus signatures are not independent variables.

Despite these shortcomings, the first arm of OCTOPUS has demonstrated that it is possible to run efficient randomised phase II trials using a platform trial design. We also showed that it is possible to make study-entry biopsies mandatory in these studies where technically feasible without compromising recruitment – 140 patients were recruited in 24 months, with study-entry biopsies obtained in 64% of participants.

In summary, the first randomised trial of a dual mTORC1/2 inhibitor in ovarian cancer showed that the addition of vistusertib to weekly paclitaxel is safe and achievable in recurrent platinum-resistant/refractory ovarian high grade serous carcinoma but does not improve PFS or OS in an unselected population. Potential predictive biomarkers will need to be evaluated in separate study cohorts.

## Data Availability

All data produced in the present study are available upon reasonable request to the authors

## Acknowledgements

We thank all the patients who agreed to participate in this study; all participating sites, principal investigators and site staff; Cancer Research UK Glasgow Clinical Trials Unit, AstraZeneca, NIHR CRN Cancer Alliance, NHS Greater Glasgow and Clyde Sponsor Pharmacy Team, Translational Pharmacology Laboratory, WWRC, University of Glasgow, Lady Garden Foundation, European Society of Medical Oncology (GG recipient of translational research fellowship). the NIHR Biomedical Research Centres at The Royal Marsden NHS Foundation Trust/Institute of Cancer Research and Imperial, the CRUK/NIHR Experimental Cancer Medicine Centres at Imperial and The Institute of Cancer Research.

---

**Table S1.**
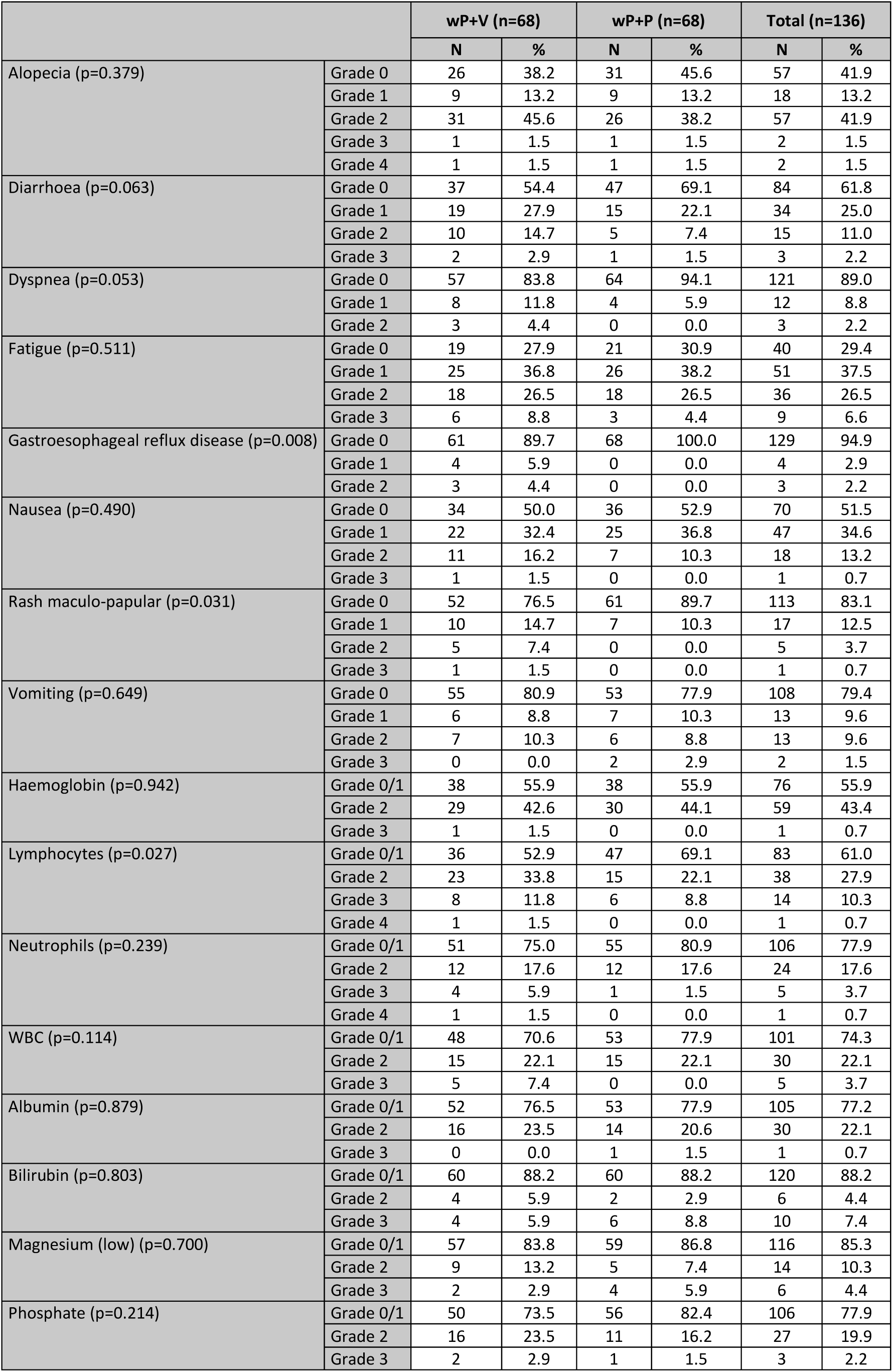
Adverse events that occurred during combination therapy at grade 2 or above in >10% of patients in either arm or that are different significantly between the arms

**Table S2.** Treatment delivery of weekly paclitaxel and vistusertib/placebo

| Treatment |  |  | wP+V (n=68) | wP+P (n=68) | Total (n=136) |
| --- | --- | --- | --- | --- | --- |
| Paclitaxel | Dose intensity | Median (Standard deviation) | 97.7 (12.1) | 97.9 (13.9) | 97.8 (13.0) |
|  | Number of dose reductions | 0 | 50 (73.5%) | 54 (79.4%) | 104 (76.5%) |
|  |  | 1 | 14 (20.6%) | 6 (8.8%) | 20 (14.7%) |
|  |  | 2 | 3 (4.4%) | 4 (5.9%) | 7 (5.1%) |
|  |  | 3 | 1 (1.5%) | 4 (5.9%) | 5 (3.7%) |
|  | Cycle delays / omissions | 0 | 16 (23.5%) | 16 (23.5%) | 32 (23.5%) |
|  |  | 1 | 20 (29.4%) | 17 (25.0%) | 37 (27.2%) |
|  |  | 2 | 11 (16.2%) | 17 (25.0%) | 28 (20.6%) |
|  |  | 3 | 8 (11.8%) | 11 (16.2%) | 19 (14.0%) |
|  |  | 4+ | 13 (19.1%) | 7 (10.3%) | 20 (14.7%) |
| Vistusertib / placebo | Dose intensity | Median (Standard deviation) | 99.7 (14.0) | 100.0 (12.4) | 99.9 (13.2) |
|  | Number of dose reductions | 0 | 41 (60.3%) | 58 (85.3%) | 99 (72.8%) |
|  |  | 1 | 14 (20.6%) | 6 (8.8%) | 20 (14.7) |
|  |  | 2 | 9 (13.2%) | 4 (5.9%) | 13 (9.6%) |
|  |  | 3 | 4 (5.9%) | 0 (0.0%) | 4 (2.9%) |
|  | Missed daily doses | Yes | 43 (63.2%) | 24 (35.3%) | 67 (49.3%) |

**Figure S1.**
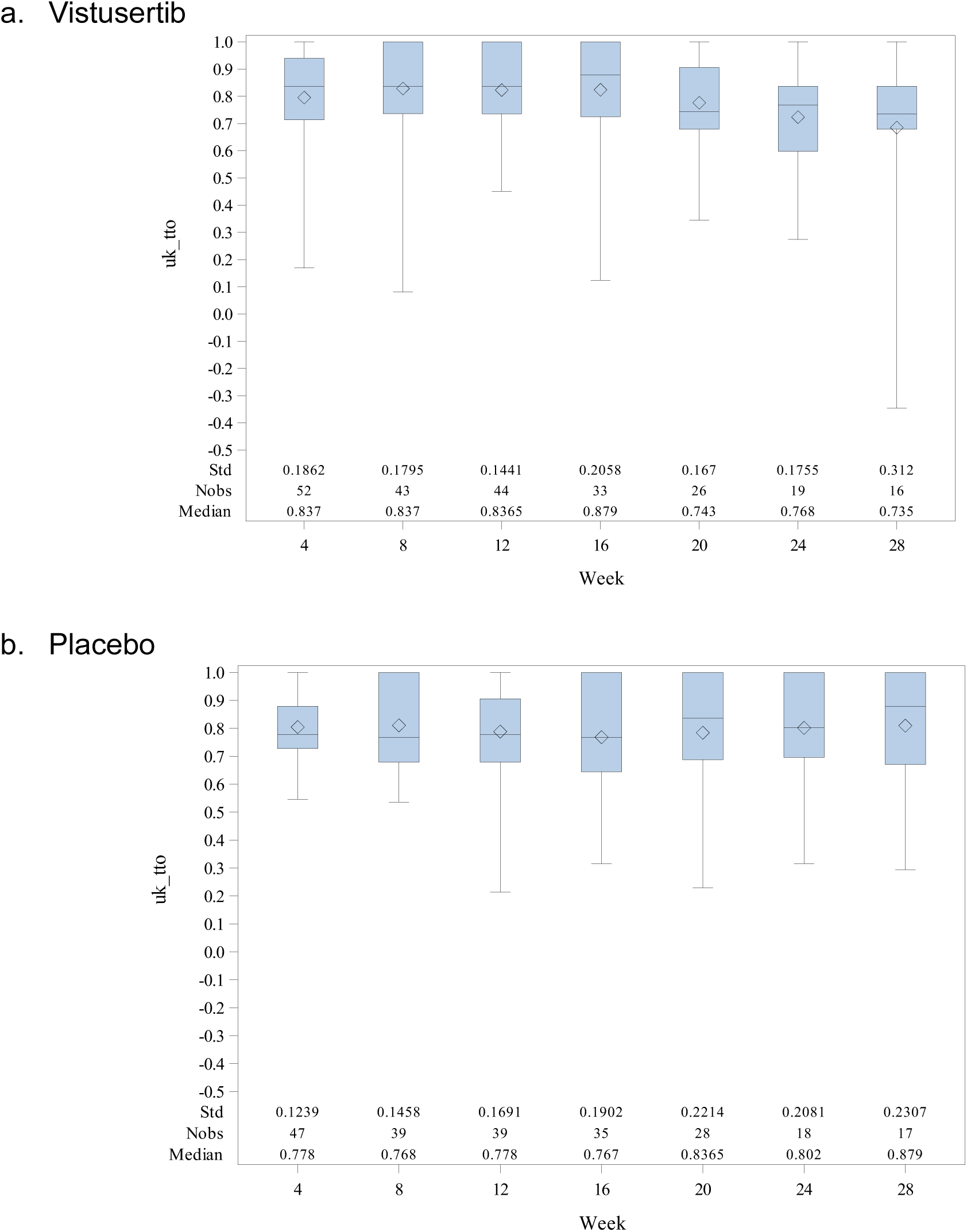
EQ5D scores by time-point.

**Figure S2.**
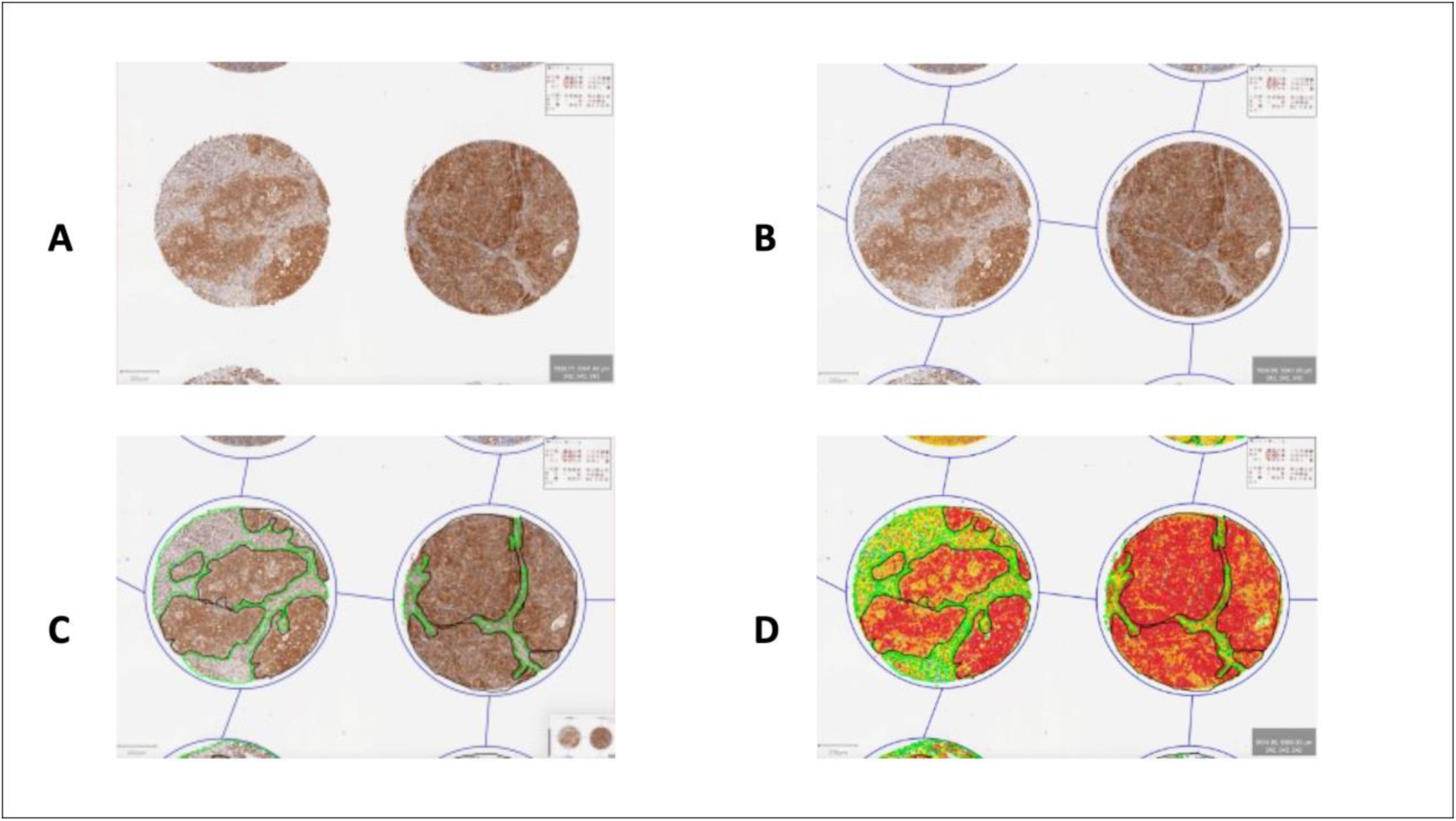
Hierarchical pathway on QuPath for PTEN Scoring.

**Figure S3.**
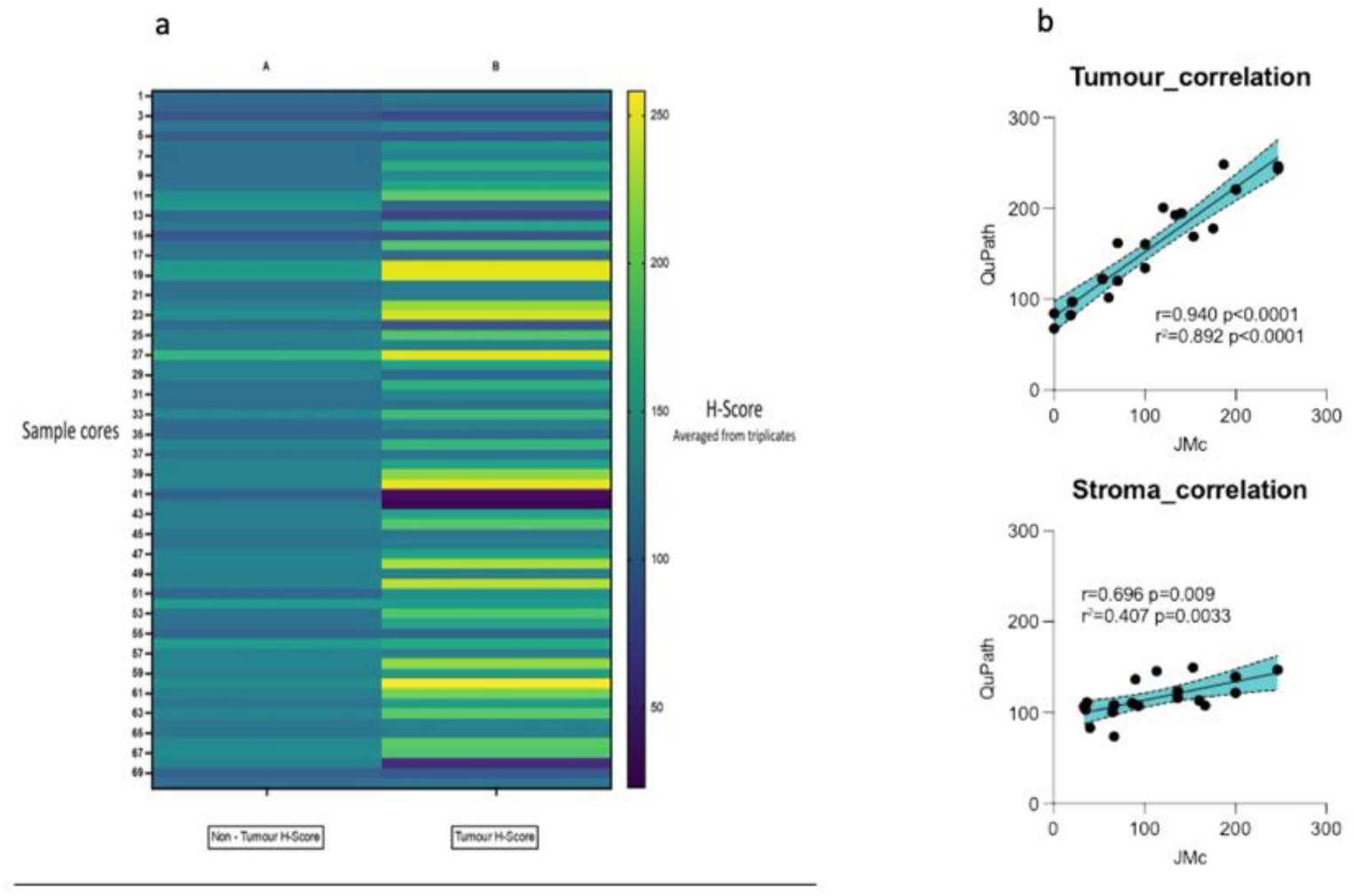
Quantitative PTEN IHC a: heatmap with average H-score in tumour and non-tumour cells across the TMAs; b: correlation between QuPath and pathologist scores (r=0.94, p<0.0001 for tumour: r=0.70, p=0.009 for non-tumour).

**Figure S4:**
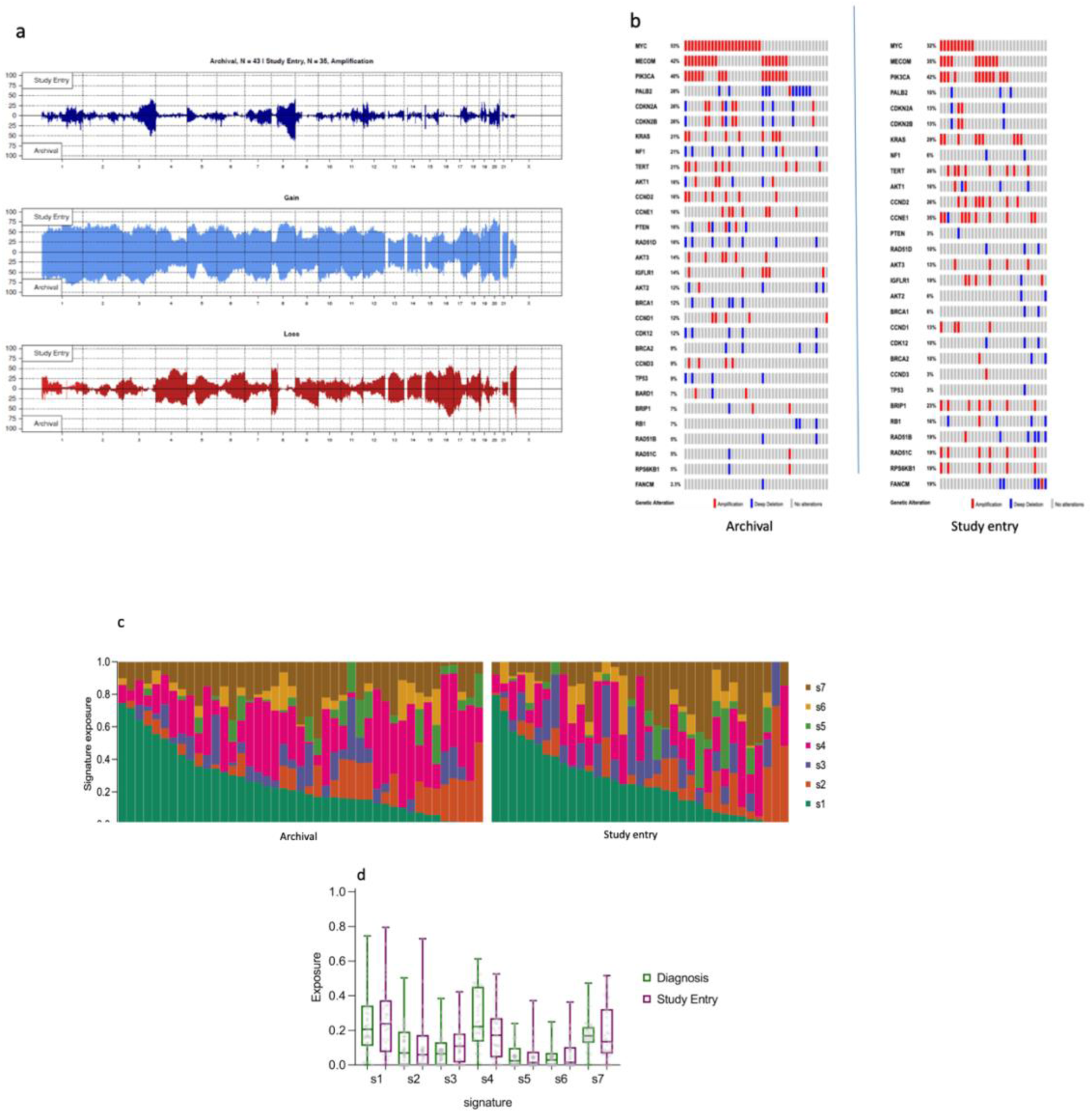
Copy number analysis a: Global amplifications, gains and losses in the whole cohort in the two cohorts; b: Focal somatic copy number alterations in 18 key genes in archival (left) and study entry biopsies (right); c: Copy number (CN) signature exposures in archival and study entry samples (each column represents one sample); d: CN signature exposure in study entry and archival cohorts. There was no statistical difference in CN exposures across the two groups.

## Supplementary Methods

### Immunohistochemistry and PTEN expression

Triplicate 1mm cores were obtained from 70 archival samples (68 patients) and a TMA was constructed and stained for PTEN (Cell Signaling #9559, 1:250) after deparaffinization and rehydration. Antigen retrieval was performed by microwave, pH 6.0 citrate buffer. Secondary antibody was vector rabbit ABC #PK6101. Negative control was performed using vector rabbit IgG serum. The PTEN antibody was validated using PC3 and 22RV1 xenografts. PTEN expression was initially quantified by two experts in pathology. Slides were scanned (Aperio ScanScope scanner) and Aperio.svs DAB images imported into QuPath (version v0.3.0) from a secure networked server. Following TMA de-arraying, cores were adjusted to 1.2mm diameter and tissue folds removed.

Cells were detected using the haematoxylin channel using default parameters. Cell type classification was conducted using training annotations (non-tumour and tumour) to train a random forest classifier to distinguish tumour and non-tumour compartments, under the direction of pathologists. We evaluated PTEN staining in both in tumour and non-tumour cells and the intensity was described as negative (0), weak (1+), moderate (2+) and strong (3+) according to these predefined DAB thresholds (negative = <0.05; 1+ = 0.05, 2+ = 0.15, 3+ = 0.4). Histoscore (H-score) was used as an output and calculated as follows: [(0 x % negative cells) (1 × % 1+ cells) + (2 × % 2+ cells) + (3 × % 3+ cells)], with a range of 0-300. We validated our PTEN scoring by randomly assigning 20% of the cores to be scored semi-quantitively by an experienced pathologist. The triplicates of each TMA sample were concordant in terms of tumour cellularity (p<0.001 using one-way ANOVA) and there was no difference in the intensity of PTEN staining if evaluated in the whole cells, in the cytoplasm or only in the nucleus of tumour and non-tumour cells. Whole cell scores were used to assign the final H-score for both tumour and non-tumour areas. In four cases, one triplicates was not assessable while for three samples only one core each was scorable. In two cases, more than one sample per patient was available; one and we randomly selected one of them. We calculated median tumour and non-tumour H-Score for each sample and compared median non tumour H-score with the median tumour H-score. We pre-defined PTEN high as median tumour H-score ≥ corresponding median non tumour H-score. PTEN low was pre-defined as median tumour H-score < corresponding median non-tumour H-score (1, 2).

### Quality check of samples, DNA extraction and sequencing

From each FFPE block, an H&E slide was prepared, and all samples underwent pathological review (two researchers independently performed a blinded review of each representative H&E and then all the cases were reviewed by a pathologist with expertise in Gynaecological malignancies). All samples confirmed as ovarian high grade serous carcinoma (HGSC) and with a tumour cellularity >20% were marked up and sections were cut in the Experimental Cancer Medicine Centre (ECMC)/Imperial Cancer Biomarker Resource Centre (ICBRC) laboratory.

DNA was thereafter extracted from 5–10 consecutive 10 μm sections using QIAmp DNA FFPE Tissue Kit (Qiagen, UK) according to the manufacturer’s protocol. 50-200ng was sheared with a Covaris LE220 focused ultrasonicator (Covaris, Woburn, MA) to produce 100-200bp fragments. Quantity was assessed using Qbit and they were immediately frozen in an ultra-low temperature freezer with a remote alarm system. Thereafter libraries were prepared in batches.

### Shallow Whole Genome Sequencing

sWGS was executed using a HiSeq4000 system (Illumina Cambridge, UK), with paired-end 150 bp protocols. The input DNA was 250 ng according to the manufacturer’s instructions. Assuming a mean ploidy of 3 and 30kb bin size, we aimed to get 10 million reads per sample. This should allow detection of a CN change of 1 (tumour purity 0.6) – 2 (tumour purity 0.3) at 80% power, alpha=0.01 https://gmacintyre.shinyapps.io/sWGS_power/. Five 150PE HiSeq4000 lanes allowed us to sequence 146 DNA samples. The minimum number of reads per sample was set at 5–10 million (mean coverage of 0.1×).

### Absolute copy number and copy number signature calling

sWGS reads were aligned to reference human genome hg19 using the BWA-MEM (3). Relative copy numbers (CN) were obtained for predefined 30kb bins using a modified version of the QDNASeq package (4). We obtained absolute CN using ACE (5). For each sample, we looked at top 10 best fits for purity and ploidy from ACE and chose the best fit that closely matched the *TP53* Variant Allele Frequency, as determined using targeted next-generation sequencing (Illumina AmpliSeq). In most cases, the ACE top fit was found to be the closest to *TP53* VAF. We defined a CN<2 as loss, 2-2.5 as normal, CN ≥2.5 but <5.0 as gain and ≥5 as amplification. CN signatures were called as previously described (6) and compared between cohorts using Kruskal Wallis test with Dunn’s multiple comparisons test on pairwise analysis of signatures-by-cohort.

### Statistics and analyses

CN signature data are compositional (i.e. they sum to 1 in each sample). Thus, we selected one normalizing signature to perform a COX regression analysis. Signature 5 was chosen as it had the lowest variability across our two cohorts (archival and study entry). To avoid division errors, all signature exposures equal to 0 were converted to 0.01. The normalization of the remaining signature exposures was calculated using the log ratio of their exposure to the exposure of signature 5. A p value of 0.05 or lower was considered statistically significant. Unless otherwise specified, statistical analyses were performed using IBM SPSS Statistics (v27).

